# Continuous bedside neuroimaging using high-density diffuse optical tomography in a pediatric patient on extracorporeal support

**DOI:** 10.1101/2024.08.12.24311750

**Authors:** Sophia R. McMorrow, Sung Min Park, Tessa G. George, Chloe M. Sobolewski, Dalin Yang, Kelsey T. King, Jeanette Kenley, Christopher D. Smyser, Joseph P. Culver, Kristin P. Guilliams, Ahmed S. Said, Adam T. Eggebrecht

## Abstract

**Background:** Extracorporeal membrane oxygenation (ECMO) provides life support for severe, reversible cardiac or respiratory failure, yet is associated with significant neurological risks including stroke. Currently available neuroimaging methods have limited sensitivity or specificity to detect early brain injury, have little real-time ability to assess interventions, and/or pose additional risks. Here, we present a case study of high-density diffuse optical tomography (HD-DOT) for bedside neuroimaging in pediatric ECMO.

**Methods:** A young infant supported on ECMO following cardiac surgery underwent continuous HD-DOT imaging over a two-hour duration that included baseline support and a clamp trial to test the ability to separate from ECMO. After stringent data quality assessments, we estimated cortical parcel-based brain functional connectivity (FC), evaluated spatial correlations between neighboring temporal epochs throughout the recording to evaluate test-retest reliability of brain FC, and calculated paired t-tests between the brain-wide set of test-retest values to test for significant changes in brain FC.

**Results:** High-fidelity bedside HD-DOT data were acquired without disruptions to patient care. During the baseline period, we observed strong test-retest with consistent bilateral FC patterns. Significant disruptions in cortical FC reflected concurrent changes in cerebral blood flow during the clamp trial and persisted after ECMO resumed.

**Conclusions:** Our results demonstrate the feasibility of continuous bedside HD-DOT neuroimaging in pediatric ECMO. HD-DOT can potentially provide clinically relevant information on cortical FC during ECMO support.

## INTRODUCTION

Extracorporeal membrane oxygenation (ECMO) provides life-saving support for severe cardio-pulmonary failure refractory to conventional therapies^1^. Pediatric ECMO use has increased since its first successful use^2^, yet ECMO-associated neurological morbidities, including stroke, persist^3^.

Early brain injury detection is imperative for neuroprotective interventions and potentially improving neurodevelopmental outcomes, but capabilities are limited. Head ultrasound (HUS) and computed tomography (CT) are bedside deployable, but only assess for major structural changes and have low predictive validity of neurological injury^4^. Magnetic resonance imaging (MRI) is typically not compatible with ECMO circuitry. Electroencephalography (EEG) provides data on brain electrical activity, reflecting synchronous neuronal activity from the cerebral cortex, but prolonged EEG can cause skin breakdown^5^.

Optical imaging based on functional near-infrared spectroscopy (fNIRS), which assesses relative changes in hemoglobin concentrations, has promise as a bedside neuroimaging tool^6^. Recent developments in high-density diffuse optical tomography (HD-DOT), utilizing dense sets of overlapping multi-distance fNIRS measurements, provide image quality comparable to functional MRI (fMRI)^7^. Previous HD-DOT studies show that low-frequency temporal correlations in cortical hemodynamics– termed functional connectivity (FC)–are sensitive to altered physiology due to stroke^8,9^ and seizures^10^. We aimed to assess the feasibility of HD-DOT for continuous, bedside neuroimaging of cortical FC in a pediatric ECMO patient.

## METHODS

### Data Availability

Data are available from the corresponding author upon request.

### Patient characteristics

A young infant with hypoplastic left heart syndrome was supported on veno-arterial ECMO after cardiac surgery. Additional clinical information is available in the Supplementary Materials. Consent for the study and potential publication of any research findings was obtained from the legal representative of the patient and the study was approved by the Institutional Review Board of Washington University in St. Louis School of Medicine.

### Data Collection

The HD-DOT system provides volumetrically reconstructed spatially distributed measurements of relative changes in oxygenated (HbO), deoxygenated (HbR), total (HbT=HbO+HbR), and difference hemoglobin (HbD=HbO-HbR) in both cortical and superficial tissues (scalp and skull). Details of the HD-DOT instrument and analyses are previously described^11^ with additional information available in the Supplementary Materials. Standard physiological variables collected included heart rate, mean arterial pressure, oxygen saturation, renal near-infrared spectroscopy, end-tidal carbon dioxide, and ECMO parameters (e.g., ECMO flow rate). Data presented focus on HD-DOT data collected continuously for over 2.5 hours on the third ECMO day, including a clamp trial to assess the ability to discontinue ECMO.

### Functional Connectivity Analyses

HD-DOT data were bandpass filtered (0.009-0.08 Hz) prior to volumetric reconstruction. We calculated FC as zero-lag temporal Pearson correlations between the mean time trace within a given cortical parcel and all cortical nodes within the HD-DOT field of view. These FC maps were generated for all cortical parcels and for each 5-minute epoch throughout the recoding.

### Statistical Analyses

To evaluate test-retest reliability of brain FC, we calculated spatial correlations between parcel-matched FC maps in neighboring temporal epochs throughout the recording. To test for significant changes in brain FC relative to the ECMO course, we calculated paired t-tests between the brain-wide set of test-retest values for each pair of neighboring epochs.

## RESULTS

The HD-DOT console measured the patient’s superficial and cortical hemodynamics via a custom cap (**Figure 1**), and recorded data continuously for 2.5 hours (**Figure 2**). Multiple real time data quality assessments ensured strong and stable coupling between the HD-DOT cap and scalp (**Supplementary Figure 1**). Throughout the baseline period (first 90 minutes), maps of low frequency temporal correlation exhibited consistent bilateral correlations (e.g., parcels in left motor or left visual cortex for HbO, HbR, and HbT is shown in **Figure 3**). During the clamp trial, strongly altered temporal correlations were apparent with cessation of ECMO flow. Following the clamp trial, the motor and visual parcels showed persistent right-hemisphere-wide disruption in HbO and HbR, suggesting asymmetric recovery of cerebral oxygen delivery, despite return of the physiological variables to baseline (**Supplementary Figure 2**). Stability quantification of FC maps using test-retest analyses for quiescent epochs revealed consistent spatial patterns of variability with strong and significant disruptions during the clamp trial (**Figure 3C-D**). Brain MRI completed three months later was without evidence of chronic infarct.

**Figure 1.**
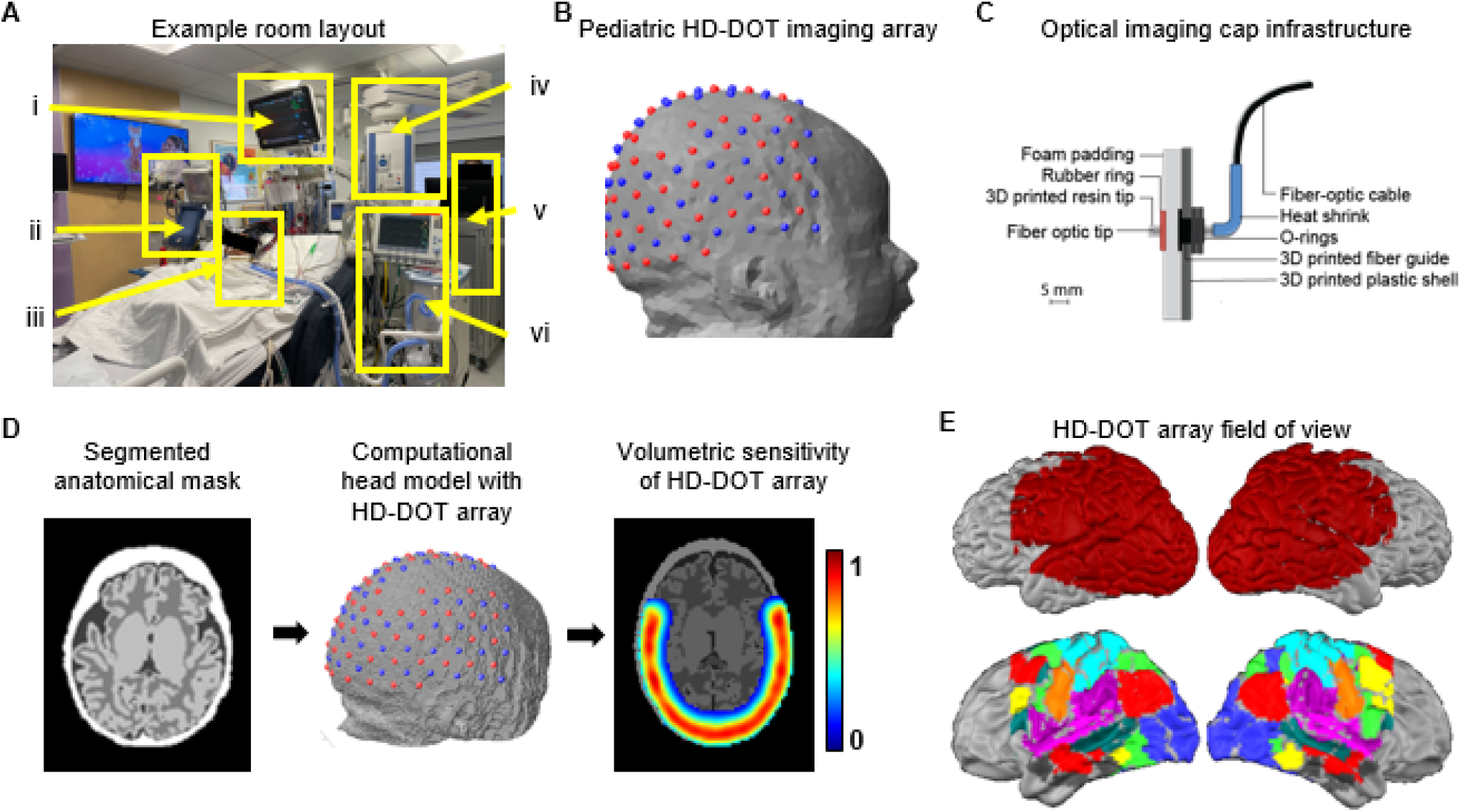
Bedside neuroimaging with HD-DOT. **A** HD-DOT system deployed in a critical care setting, including i) vital sign monitor, ii) ECMO circuit, iii) patient in bed wearing HD-DOT cap, iv) support equipment, v) HD-DOT console, and vi) ventilator. **B** Pediatric HD-DOT imaging array on representative infant head. **C** 3D-printed TPU-based optical imaging cap provides structural support and couples the scalp to optical fibers (whose tips are covered with transparent resin). **D** Age-appropriate atlas MRI is combined with source (red) and detector (blue) locations to computationally model the HD-DOT sensitivity on the patient anatomy. **E** Field of view on cortical surface model. Data registration and standardization aligns patient data with common atlases for population-referenced analyses. Here, we show the Gordon parcellation, colored by functional network, intersecting the HD-DOT field of view.

**Figure 2.**
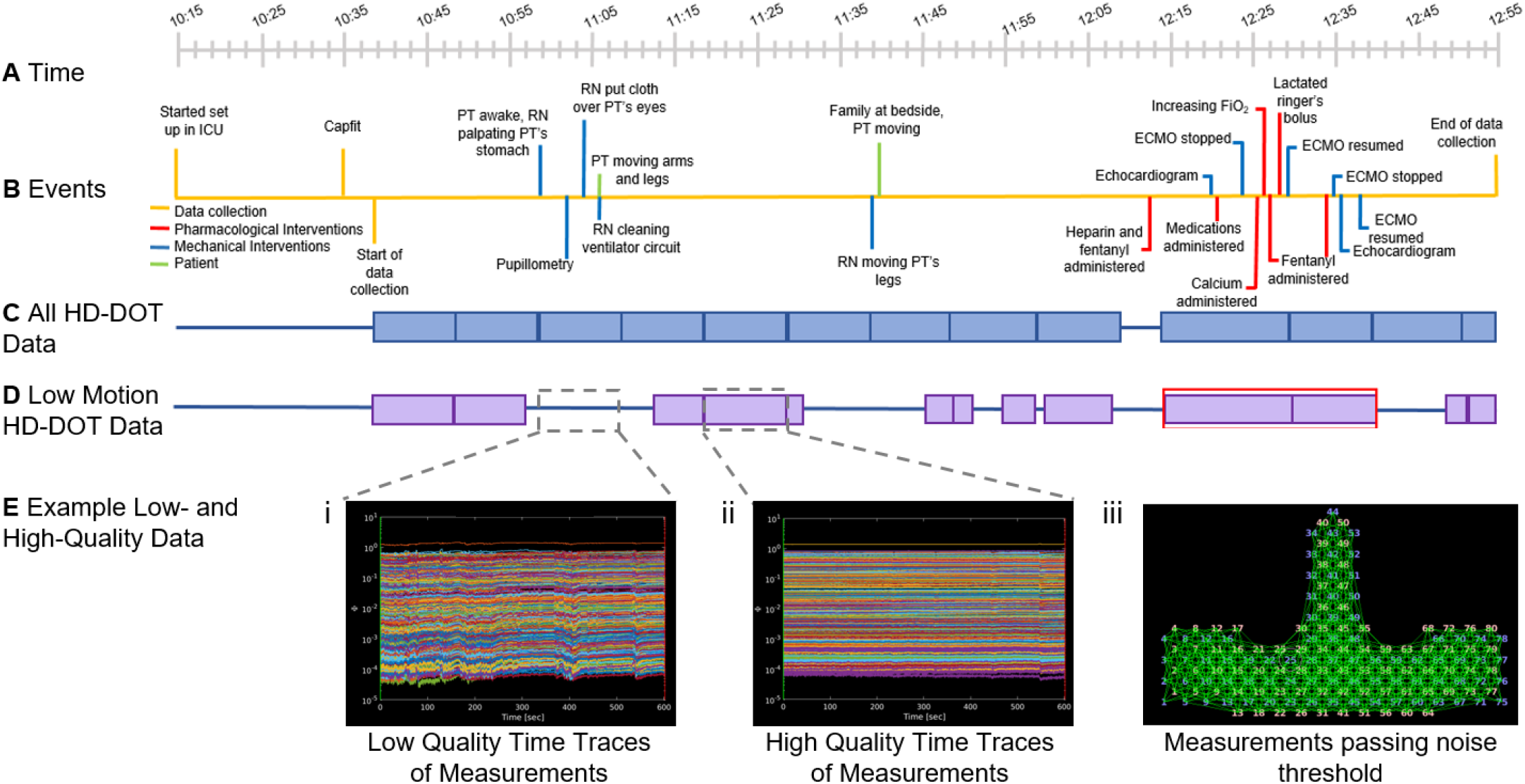
Data Collection and Data Quality Assessment. Timecourse of data collection including: **A** Time of day; **B** Events during data collection; **C** All HD-DOT data collected during this session; **D** Quiescent HD-DOT data. Red box: period of interest including a clamp trial. **E** Example data quality metrics. i-ii) Time traces of all good 830nm HD-DOT measurements. i) Noisy traces indicate disruptions in optode coupling. ii) Smooth time traces indicate stable and robust optode coupling. iii) Measurement channels with a standard deviation lower than 7.5% of the mean signal are shown as green lines between optodes on a flattened two-dimensional representation of the HD-DOT array.

**Figure 3.**
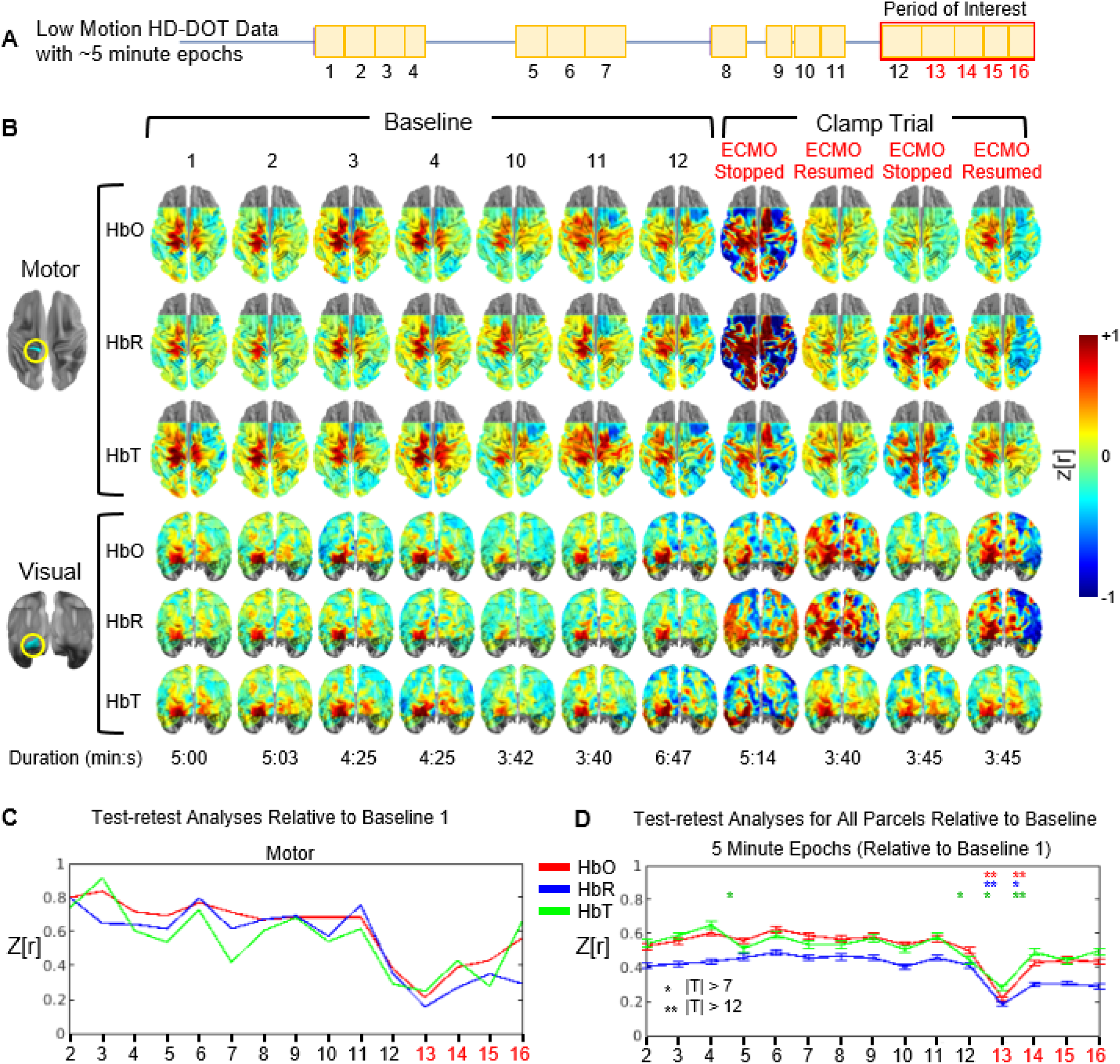
Longitudinal mapping of functional connectivity. **A** Low motion HD-DOT data divided into approximately 5-minute periods. Red text denotes clamp trial epochs. **B** Fisher’s z-transformed Pearson correlation maps for parcels in the motor and visual cortices show consistent bilateral correlation at baseline and disruptions during the clamp trial. **C** Test-retest analyses for a motor parcel and **D** for all parcels relative to an initial baseline period show consistent FC patterns at baseline and disruptions during the clamp trial. Error bars show standard error of the mean. *P* values were calculated with a paired samples *t*-test, \**p* < 1^-10^, \*\**p* < 1^-20^.

## DISCUSSION

Current neuroimaging tools do not provide bedside evaluation of spatial-temporal FC variability. Herein, we assessed cortical FC via bedside HD-DOT on ECMO for over two hours, including during a clinical clamp trial. To our knowledge, FC has not been previously investigated in ECMO patients. While FC does not provide flow quantitation, its clear sensitivity to alterations of cerebral oxygen delivery highlights its potential as a novel bedside neuroimaging tool. In this neonatal patient, FC maps exhibited consistent patterns of correlation throughout the baseline period, including strong bilateral connectivity patterns in representative motor and visual regions. These findings are consistent with infant MRI literature, where healthy infants show bilateral correlation in homotopic regions, with interhemispheric FC in the primary sensorimotor and visual regions present at birth^12–14^. Additionally, these consistent correlation patterns were disrupted during the clamp trial, accompanied by large fluctuations in physiological variables. Considering that this patient was deemed not ready to separate from ECMO, this FC disruption may be an additional indicator that the patient’s native cardiopulmonary function alone did not provide adequate cerebral oxygen delivery. Further, our results suggest that FC remained disrupted immediately after ECMO resumed, possibly caused by hemodynamic stress incurred during the clamp trial.

### Clinical Implications and Future Directions

Our methods provide the first demonstration of fMRI-comparable data at the bedside in this high-risk patient population. Further tool development may provide actionable information in real time.

### Limitations

If a patient is not ready to stop ECMO, an unsuccessful clamp trial presents a significant alteration in global oxygen delivery; alterations in FC due to focal blood flow disruptions, such as during a small thromboembolic stroke, are unknown. Future studies are needed to examine FC patterns during ECMO support and short- and long-term outcomes of disruptions. Additionally, the field of view of the HD-DOT system lacks full coverage in the prefrontal area, despite presenting a substantial increase in the field of view compared to prior bedside infant HD-DOT systems^11^.

## Conclusions

Our preliminary findings establish the feasibility of bedside FC neuroimaging with HD-DOT during ECMO, a population at high risk for stroke. ECMO patients are a complex, heterogeneous population, and further studies are necessary to understand the short- and long-term impact of FC variability during ECMO on outcomes.

## Data Availability

All data produced in the present study are available upon reasonable request to the authors

