## Supplementary Information for "Continuous bedside neuroimaging using high-density diffuse optical tomography in a pediatric patient on extracorporeal support"

### Supplementary Materials

#### *Patient Characteristics*

A full-term-born female neonatal patient was postnatally diagnosed with hypoplastic left heart syndrome with mitral and aortic atresia after presenting to the pediatrician's office in cardiogenic shock with closing of the patent ductus arteriosus. She was stabilized in the cardiac intensive care unit and underwent the Norwood procedure with a right ventricle-pulmonary artery shunt. As she was unable to safely separate from bypass following the procedure, the infant was centrally cannulated for veno-arterial extracorporeal membrane oxygenation (ECMO) in the operating room before returning to the cardiac intensive care unit. The patient had head ultrasounds prior to ECMO and daily while on ECMO support. No acute intracranial abnormalities were shown on any ultrasound. The patient was monitored with EEG for the first 48 hours of ECMO support, and no seizures or abnormalities were detected.

#### *HD-DOT Data Acquisition and Analyses*

##### *HD-DOT System*

We used a continuous-wave HD-DOT system optimized for pediatric patients with an optode array consisting of 80 dual-wavelength laser diode sources (685nm and 830nm) and 78 avalanche-photodiode (APD) detectors. The HD-DOT console is bedside-deployable (**Figure 1A**), and contains opto-electronics similar to those previously described[1] including five source boxes, 16 detector boxes, and five Focusrite analogue-to-digital converters. The HD-DOT system collected data at a 10 Hz frame rate.

The sources and detectors were coupled to the head via six-meter-long optical fibers attached with a 3D-printed imaging cap and arranged in an interdigitating array covering bilateral parietal, occipital, temporal, and prefrontal areas (**Figure 1B**). This arrangement supported over 1200 overlapping, multi-distance source-detector pair measurements (first through fourth nearest neighbors at 11 mm, 25 mm, 33 mm, and 40 mm, respectively). The HD-DOT cap was designed to optimize patient comfort during extended scanning sessions (**Figure 1B,C**). The 3D-printed cap shape was modeled off a representative anatomical T1-weighted MRI to provide an infant head shape, printed using thermoplastic polyurethane (TPU) to provide flexible structural support, and included soft foam padding on the inside to minimize pressure on the head and ensure patient comfort. To enhance flexibility and accommodate head circumferences ranging from 34-39 cm, the cap includes cutouts on the top and adjustable Velcro straps. Top-hat style spacers were 3D printed using polylactic acid fiber and allow moderate radial movement (perpendicular to the scalp surface) of the fiber tips. Right-angle optical fibers allow the cap to be supported on the hospital bed for comfortable extended scanning. 3D-printed transparent resin tips were added to the fibers and secured with heat shrink to defocus light and increase the size of the optical tip footprint for comfort, while still allowing the tip to comb through hair and sit at a 90-degree angle against the scalp. Compressible rubber rings secure the optodes to maintain adequate coupling to the scalp and patient comfort. The optical fiber tip protrudes 3 mm through the foam to facilitate combing through the hair. Flexible optical fiber bundles allow the cap to be placed up to six meters from the console.

***Anatomical Light Modeling, Data Preprocessing, Image Reconstruction, and Spatial Normalization***

An age-appropriate anatomical atlas and segmented mask[2–4] were used to model light propagation and depth sensitivity of the HD-DOT system using the NeuroDOT preprocessing pipelines in MATLAB (**Figure 1D,E**). The segmented mask was created from T1- and T2-weighted MRIs from an infant with comparable head circumference and age as the patient. A computational head mesh was generated from the segmented mask and combined with the optode array placement based on photos taken during data collection. Colored markers on the cap and subject-specific facial landmarks were used to ensure alignment of anatomy with the HD-DOT array. The head model was used to model light propagation and depth sensitivity of the HD-DOT array using the NIRFASTer light modeling software[5] (**Figure 1D**). The field of view was established as head tissue with a normalized flat field reconstruction greater than 10%.

Quiescent epochs of data were retained for further analyses where noisy segments of data were cropped manually based on data-quality metrics of time traces. Specifically, source-detector measurements with a temporal standard deviation less than 7.5% of the mean signal were retained for further analyses. General pre-processing, image reconstruction, and spectroscopic unmixing procedures for HD-DOT have been previously described[1,6]. The mean across all first nearest neighbor measurements was constructed as an estimate of superficial tissue signals and was regressed from all measurements before volumetric reconstruction. Volumetrically reconstructed data were spatially smoothed (3D Gaussian kernel, 3 mm full width at half maximum) and down-sampled to 1 Hz. The volumetric data were spatially registered from the neonatal model anatomy to the standard MNI-based atlas space using an affine transform for standardized analyses. The atlas-registered HD-DOT data were intersected with publically available atlas-based Gordon cortical parcels[7] to provide a direct referencing to common putative functional networks (**Figure 1E**).

***Data Acquisition***

Prior to data collection, the console was positioned in the room such that it was out of the way of any monitors, ventilators, and the ECMO circuit to minimize disruption to the medical staff's workflow (**Figure 1A**). The six-meter long, flexible optical fiber bundle was fastened to the hospital bed and other available supports (e.g., table, console). The weight of the cap was supported by the hospital bed, and healthcare professionals gently raised the patient's head to position it in the cap, then secured the cap with Velcro straps. The patient's head and neck were cushioned with blankets and supported with a pillow. The cap was positioned such that the bottom row of optodes rested just above the ears (**Figure 1B**). Photographs at different angles were collected after the cap was placed on the patient for data registration used in image reconstruction.

To optimize data quality, multiple real-time metrics were assessed during cap placement, including the temporal mean light level for each source-detector pair plotted as a function of source-detector distance (**Supplementary Figure 1A**), the mean light level for each optode to ensure adequate coupling of the optical fiber with the scalp (**Supplementary Figure 1B**), and the signal to noise ratio (SNR) of the pulse signal at each optode position (**Supplementary Figure 1C**). In an ideal cap fit, the temporal mean of measurements across the cap decay with a log-linear fall-off related to the distance between source-detector pairs, a uniform distribution of coupling quality throughout the entire cap, and a similarly strong and consistent pulse SNR across the cap.

A total of 3 hours and 15 minutes of data were collected during the third and fourth days of ECMO support. For simplicity, we present the results from the first day of data collection, which lasted a total of 2 hours and 10 minutes (**Figure 2**). This scan session included a 90-minute period of relative inactivity, during which healthcare personnel performed routine clinical care, such as examining the ECMO circuit, suctioning the patient's airway to remove secretions, and adjusting the patient's position.

Towards the end of this scanning session, a clamp trial was conducted to assess the patient's ability to be weaned from ECMO support. To prepare for the clamp trial, healthcare personnel modified the flow rate of various medications, including calcium, dexmedetomidine, epinephrine, milrinone, morphine, and nitroprusside. During the clamp trial, ECMO flow was stopped for five minutes. While ECMO was stopped, $SpO_2$, MAP, and renal NIRS decreased (**Supplementary Figure 2**). Notably, $SpO_2$ and MAP dropped to lows of 61% and 29 mmHg, respectively. To prevent blood clots, ECMO was briefly resumed for about four minutes, during which $SpO_2$, MAP, and renal NIRs returned toward baseline levels (**Supplementary Figure 2**). Following this, ECMO was stopped a second time for about four minutes, causing $SpO_2$, MAP, and renal NIRS to decrease again. However, it is important to note that a decrease in $SpO_2$ was expected and considered tolerable for this patient, who had single-ventricle physiology. Patients with single ventricle physiology have lower peripheral $SpO_2$ values varying from 75-85%, while $SpO_2$ for healthy patients is ≥96%[8]. While monitoring vitals during the clamp trial, physicians determined that the patient was not yet ready to come off ECMO, so the clamp trial ended and ECMO resumed. The total duration of the clamp trial was 13 minutes. Approximately 20 minutes of data were collected after the clamp trial. Of the 2 hours and 10 minutes of data collected during this scan session, 1 hour and 15 minutes of quiescent data were retained for further analyses (**Figure 2**).

***Offline Data Quality Assessment***

Raw data quality metrics were also analyzed after data collection using the NeuroDOT software to identify high-quality and low-motion data for further analysis (https://www.nitrc.org/projects/neurodot). Time traces of channel measurements during data collection were visualized (**Figure 2Ei-ii**). Measurement channels with a standard deviation lower than 7.5% of the mean signal are shown as green lines between optodes (**Figure 2Eiii**). Noisy measurement channels above this threshold are removed. Smooth time traces indicate stable and robust optode coupling, while noisy time traces indicate movement and disruptions in optode coupling.

***Systemic Oxygenation Analyses***

For analyses of high bandwidth cerebral oxygenation, prior to volumetric reconstruction, unfiltered HD-DOT measurements were converted to relative hemoglobin concentrations using the modified Beer-Lambert law and a path-length factor of six. Relative hemoglobin concentrations in superficial tissue were estimated as the mean across all non-noisy first nearest neighbor measurements. Data were aligned with concurrently recorded physiology. Bedside vitals were recorded at a frequency of 0.2 Hz and manually time-synched with HD-DOT data for analyses (**Supplementary Figure 2**).

Superficial concentrations of HbO, HbR, HbT, and HbD remained at baseline levels throughout the quiescent period prior to the clamp trial. In response to clamp trial events, HD-DOT data reveal rapid and notable changes in concentrations of HbR and HbO, and their difference HbD. When ECMO stopped, HbR immediately increased, while HbO and HbR decreased. When ECMO resumed, relative hemoglobin concentrations returned toward baseline levels seen prior to the clamp trial. When ECMO stopped the second time, HbR again immediately increased, and HbO and HbR decreased. Hemoglobin concentrations returned toward baseline levels when ECMO resumed. The response in superficial tissue to stopping ECMO flow was similar to systemic physiology; $SpO_2$, renal NIRS, and MAP immediately decreased when ECMO stopped, and returned toward baseline levels when ECMO resumed. Additionally, hemoglobin concentrations in superficial tissue have relatively smaller perturbations from baseline at 350-400 seconds and 600 seconds into the period of interest. The perturbations around 35-400 seconds coincided with the administration or modification of the flow rate of various medications. The perturbation in superficial hemoglobin concentrations at 600s coincided with the administration of calcium.

### Supplementary Figure 1

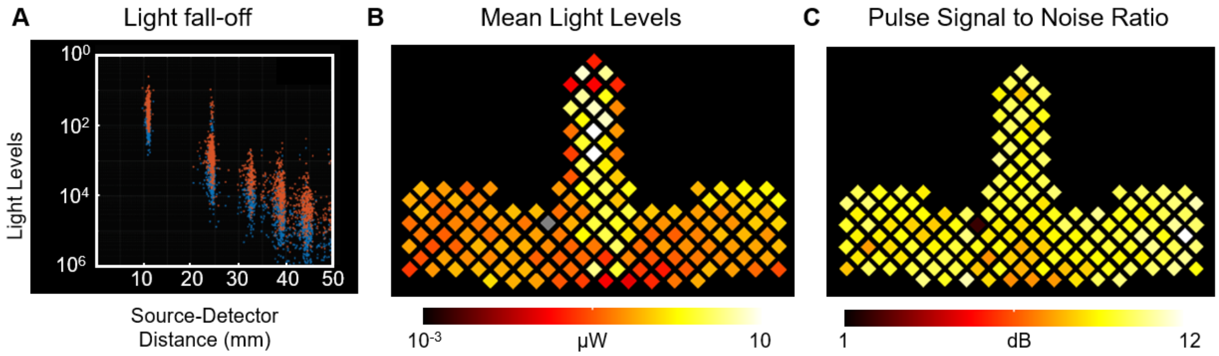


**Supplemental Figure 1 | Data Quality Assessment. A** The temporal mean of measurements across the cap decay with a log-linear fall-off related to source-detector distance. **B** The average coupling coefficients of each optode across the cap is within two orders of magnitude. **C** A high signal to noise ratio displayed on a flattened view of the cap.

### Supplementary Figure 2

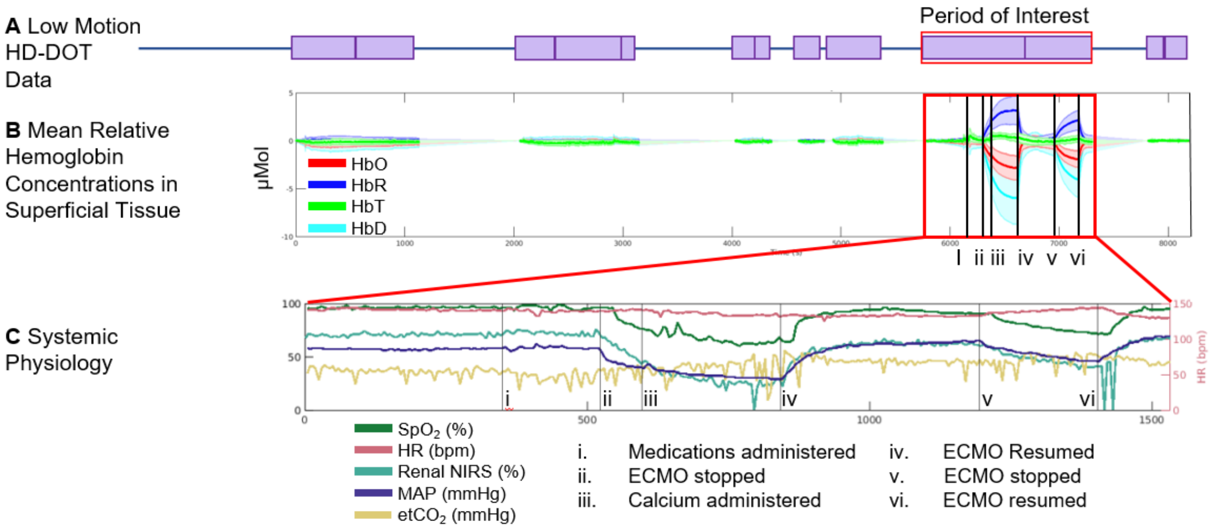


**Supplemental Figure 2 | Period of interest including a clamp trial. A** Low motion HD-DOT data. Red box: period of interest including clamp trial. **B** Mean relative hemoglobin concentrations in superficial tissue remain consistent during baseline periods, with changes induced by clamp trial events (i-vi). **C** Vitals monitored as a part of standard clinical care were obtained and aligned with HD-DOT data. $SpO_2$, MAP, and renal NIRS show immediate changes in response to clamp trial events.
